# Sentence production and sentence repetition in autistic adolescents and young adults: Linguistic sensitivity to finiteness-marking

**DOI:** 10.1101/2024.03.26.24304924

**Authors:** Teresa Girolamo, Samantha Ghali, Caroline Larson

**Affiliations:** San Diego State University; University of Kansas; University of Missouri

**Keywords:** language impairment, morphosyntax, autism, linguistic sensitivity, strategic scoring

## Abstract

**Purpose:** Despite the clinical utility of sentence production and sentence repetition to identify language impairment in autism, little is known about the extent to which these tasks are sensitive to potential dialectal variation. One promising method is strategic scoring (Oetting et al., 2016), which has good clinical utility for identifying language impairment in nonautistic school-age children across dialects of English. This report applies strategic scoring to analyze sentence repetition and sentence production in autistic adolescents and adults.

**Method:** Thirty-one diverse autistic adolescents and adults with language impairment (ALI; *n*=15) and without language impairment (ASD; *n*=16) completed the Formulated Sentences and Recalling Sentences subtests of the Clinical Evaluation of Language Fundamentals-5^th^ Ed (Wiig et al., 2013). Descriptive analyses and regression evaluated effects of scoring condition, group, and scoring condition by group on outcomes, as well as group differences in finiteness-marking across utterances and morphosyntactic structures.

**Results:** Strategic and unmodified item-level scores were essentially constant on both subtests and significantly lower in the ALI than the ASD group. Only group predicted item-level scores. Group differences were limited to: percent grammatical utterances on Formulated Sentences and percent production of overt structures combined on Sentence Repetition (ALI < ASD).

**Discussion:** Findings support the feasibility of strategic scoring for sentence production and sentence repetition to identify language impairment and indicate that potential dialectal variation in finiteness-marking did not confound outcomes in this sample. To better understand the clinical utility of strategic scoring, replication with a larger sample varying in age and comparisons with dialect-sensitive measures are needed.

---

Half of all autistic individuals are estimated to have LI, which is characterized by challenges with structural language (Boucher, 2012). LI in autism is tied to poorer educational, health, occupational and social outcomes (Howlin et al., 2013; Johnson et al., 2010; Magiati et al., 2014). Yet, autistic youth – especially if racially and ethnically minoritized – face unreliable access to speech/language services (Newman et al., 2011; Pope et al., 2022; Taylor & Henninger, 2015). One barrier to access is that given current diagnostic criteria for autism (American Psychiatric Association, 2013), schools may only assess behavior and not language in autistic students (Musgrove, 2015). Reducing disparities requires quality language assessment.

Quality language assessment requires evidence-based practice. On one hand, evidence-based practice requires reliable methods for identifying LI in autism. Finiteness-marking, or marking of tense and agreement, is one aspect of morphosyntax and a clinical marker of LI in autism (Eigsti et al., 2011; Modyanova et al., 2017); see Ash and Redmond (2014). In turn, sentence repetition and sentence production are useful for assessing morphosyntax in autism (Schaeffer et al., 2023). Evidence-based assessment also requires understanding how to interpret and use measures (Girolamo et al., 2022b; Messick, 1990). Yet, this knowledge may be inadequate. From 2004 to 2014, 75% of states disproportionately represented Black students in the disability category of speech/language impairment (Robinson & Norton, 2019). In research, studies using norm-referenced assessments in English to characterize structural language in autistic youth (ages 3-21) systematically exclude or do not account for those with LI and those who are racially and ethnically minoritized (Girolamo et al., 2023b, 2023c). Thus, the evidence base needed to inform language assessment in autism is incomplete.

A broader consideration in assessment involves the linguistic reality of the United States. Individuals of all races and ethnicities speak over 25 dialects of English (Wolfram & Schilling, 2015). Dialects vary in how they mark for finiteness, such that dialectal variation and LI each influence expressive language in terms of morphosyntactic production (Oetting et al., 2016). Yet, clinical language research assumes General American English (GAE) is the norm (Oetting, 2020), and evaluators of science may perpetuate the myth that only minoritized individuals speak dialects other than GAE (Girolamo et al. 2022a). Accurate language assessment requires both representation in research and linguistic sensitivity without racialized assumptions about language background (Plaut, 2010). This report examines sentence production and sentence repetition, focusing on finiteness-marking, in diverse autistic adolescents and adults.

## Sentence Production and Sentence Repetition in Autism

Epidemiological data support the utility of sentence repetition and sentence production for identifying LI in nonautistic youth (<u>ages 4-10)</u> (Calder et al., 2023; Klem et al., 2015). These tasks are often part of assessments, such as the Clinical Evaluation of Language Fundamentals (CELF) (Wiig et al., 2013), which are commonly used in practice and psychometrically validated (Betz et al., 2013; Nitido & Plante, 2020). While autism research does not have population-level evidence, cross-linguistic applications of these tasks support their clinical utility.

### Group Comparisons

Prior work evaluating sentence repetition and sentence production has compared groups of autistic individuals without mention of LI (ASD) and nonautistic peers. In addition, some studies compared within-autism heterogeneity: autistic individuals without LI (ASD) and autistic individuals with LI (ALI). A summary is presented in Supplementary Table 1.

In studies comparing ASD and nonautistic peers, one pattern was lower performance in ASD (Hedges’ *g* = 0.62 to 1.34). Some samples showed this pattern on both tasks using percent accuracy on the Russian Child Language Assessment Battery (ages 7-10) (Arutiunian et al., 2022; Lopukhina et al., 2019) and scaled scores on the English CELF-4 (ages 8-21) (Larson et al., 2022; Semel et al., 2003; Tyson et al., 2014). Some studies found lower scores in ASD using just sentence repetition: (a) raw scores on a Danish translation of the CELF-Preschool, 2^nd^ Ed. (ages 4-6) (Brynskov et al., 2017; Wiig et al., 2004), (b) raw scores on the Hebrew PETEL Test (ages 9-18) (Friedmann, 2000; Sukenik & Friedmann, 2018), and (c) percent accuracy on the Saudi Sentence Repetition Task (ages 5-7) (Alhassan & Marinis, 2021). In turn, scaled scores were lower in ASD (ages 7-17) on the English CELF-Revised Formulated Sentences (Landa & Goldberg, 2005; Semel et al., 1987). A second pattern was no group differences in scaled scores on the English CELF-4 Formulated Sentences and Recalling Sentences (ages 9-16) (Harper-Hill et al., 2013; Semel et al., 2003) or in percent accuracy on the LITMUS-French sentence repetition (ages 18-56) (Manenti et al., 2023; Prevost et al., 2012). A third pattern was mixed performance, with lower raw scores in ASD (ages 6-12) on sentence production but not sentence repetition using the Greek Expressive and Receptive Language Evaluation (EREL) (Georgiou & Spanoudis, 2021; Spanoudis & Pahiti, 2014). In all, findings and measurement varied.

Studies comparing ALI and ASD showed similar variation. ALI had lower percent accuracy on the Saudi Sentence Repetition Task (ages 5-7; Hedges’ *g* = 1.53) (Alhassan & Marinis, 2021) and the LITMUS-French Sentence Repetition (ages 18-56; *r* = −0.784) (Manenti et al., 2023; Prevost et al., 2012). Some studies provided only frequencies suggesting lower performance in ALI: average raw score on the English CELF-4 Recalling Sentences and Formulated Sentences (*M*_age_ = 11.0) (McGregor et al., 2012; Semel et al., 2003) and percent accuracy on the LITMUS-French Sentence Repetition (ages 6-12) (Prevost et al., 2012; Silleresi et al., 2020). A second pattern was no group differences in: (a) raw scores on the Greek EREL sentence repetition or sentence production (ages 6-12) (Georgiou & Spanoudis, 2021), (b) scaled scores on the English NEPSY Memory for Sentences (ages 7-15) (Korkman et al., 1998; Whitehouse et al., 2008), or (c) raw scores on the English CELF-3 Recalling Sentences (ages 14-15) (Riches et al., 2010). As with ASD and nonautistic comparisons, there was no one pattern.

### Limitations to Generalizability

Findings indicate sentence production and sentence repetition tasks capture linguistic heterogeneity in ALI and ASD, but there are limitations to generalizability. Like autism research overall (Russell et al., 2019), most studies selected against individuals with NVIQ < 70 and used IQ cutoffs as high as −1 *SD* (McGregor et al., 2012). Yet, estimates of NVIQ < 70 in autism are 38% to 50% (Charman et al., 2003; Loomes et al., 2017; Maenner et al., 2023). Such IQ cutoffs may fail to reflect language across the spectrum or the ways in which language and NVIQ may dissociate. In an epidemiological study of nonautistic youth with LI, the severity of LI did not differ when NVIQ was −1 to −2 *SD* versus within 1 *SD*, and only one of five language subtests differed when NVIQ was < −2 *SD* (Norbury et al., 2016). Further, except for two instances (Manenti et al., 2023; Riches et al., 2010), studies focused primarily on children, amid a need for information on autism in adulthood (Howlin & Taylor, 2015). Finally, consistent with multiple areas of autism research (Girolamo et al., 2023c; Larson et al., 2023; Steinbrenner et al., 2022; West et al., 2016), full reporting of race and ethnicity was rare (Larson et al., 2022). Hence, the utility of these tasks for diverse, older autistic individuals varying in NVIQ is unknown.

## Linguistically Sensitivity to Finiteness-Marking

Interpreting sentence production and sentence repetition from diverse autistic adolescents and adults in American English requires attention to differences in finiteness-marking due to dialectal variation (Beyer & Hudson Kam, 2012). This is not because there is a one-to-one ratio between race, ethnicity, and dialectal variation. Rather, given the linguistic reality of the United States, being linguistically sensitive through strategic scoring and examination of finiteness-marking is clinically and scientifically sound (Oetting et al., 2016). To our knowledge, this approach has yet to be used in autism research or with individuals older than ages 4 to 6 years.

### Scoring Methods for Finiteness-Marking

Recall that dialects of American English differ in how they mark for finiteness (Oetting et al., 2019; Wolfram & Schilling, 2015). In GAE, the past-tense of “eat” requires overt inflection (Wolfram & Schilling, 2015); see (1a). In African American English (AAE) and Southern White English (SWE), the past-tense of “eat” can be zero-marked, with the null marker indicating inflection; see (1b) (Wolfram & Schilling, 2015). It is not that AAE or SWE speakers would only produce (1b). Rather, (1a) and (1b) are each plausible, with differences in production rate by dialect and LI status (Cleveland & Oetting, 2013; Garrity & Oetting, 2010; Oetting & McDonald, 2001; Seymour et al., 1998).

(1) Examples of the past-tense irregular “eat” (Oetting et al., 2019)
(1a) she *ate* [overt]
(1b) she *eat*Ø [zero]
(1c) *the girl* [other]

A question, then, is how to characterize these differences in dialect, clinical status, and rate. Descriptively examining finiteness-marking can inform development of systematic approaches to scoring (Oetting & McDonald, 2001). In sentence (1), unmodified scoring only counts GAE overt forms, or (1a), as correct; any other response, including (1b) and (1c), is incorrect (Oetting et al., 2019). Hence, unmodified scoring over-identifies LI in speakers of dialects other than GAE (Hendricks & Adlof, 2017). Conversely, modified scoring is responsive to all possible instances of dialectal variation and counts (1a) and (1b) as correct (Oetting et al., 2019). In only considering other responses like (1c), as incorrect (Oetting et al., 2019), rather than production rate of overt and zero marking, modified scoring can under-identify LI (Craig et al., 2004; Hendricks & Adlof, 2017). To optimize differences across dialects, Oetting and colleagues (2019, 2016, 2021) developed strategic scoring. While strategic scoring counts (1a) and (1b) as correct, it considers the proportion of GAE and non-GAE overt forms to all overt GAE and zero forms; only other responses like (1c) are excluded (Oetting et al., 2019).

Importantly, strategic scoring has differentiated LI and typical language in 106 diverse nonautistic youth (ages 4-6) who speak AAE, GAE, and SWE when using dialect-sensitive sentence repetition and sentence production tasks. In administering a sentence repetition probe with auxiliary BE, Oetting et al. (2016) counted responses that differentiate AAE and SWE from GAE, but do not confound identification of LI, as correct: “is” for “are,” “was” for “were,” and zero third-person singular (Cleveland & Oetting, 2013; Oetting & Garrity, 2006). Strategic scoring showed high classification accuracy when considering AAE and SWE together (Se = .91, Sp = .85) and separately (AAE: Se = .89, Sp = .86; SWE: Se = .94, Sp = .83) (Oetting et al., 2016). The presence of effects of clinical status (partial η^2^ = .55, *p* < .001), but not dialect (partial η^2^ = .02, *p* = .13), on sentence repetition probe raw scores indicated dialectal variation did not confound group differences (Oetting et al., 2016).

Turning to sentence production, Oetting et al. (2019) administered four probes to nonautistic youth (ages 4-6) that targeted past-tense regular and irregular, singular and plural present auxiliary BE, singular and plural past auxiliary BE, and habitual and nonhabitual third-person singular. Across all structures, strategic scoring had higher classification accuracy than modified scoring (75% versus 66%) and more balanced sensitivity and specificity than unmodified or modified scoring (strategic: .72 and .77; unmodified: .81 and .68; modified: .51 and .81) (Oetting et al., 2019). In addition, unmodified scoring yielded differential effects of clinical group by dialect for AAE and SWE (η^2^: .27 versus .56), indicating lack of measurement invariance; conversely, strategic scoring had twice the effect of clinical group (η^2^: .38 versus .17) compared to modified scoring (Oetting et al., 2019). When separating structures, only unmodified and strategic scoring yielded differences by clinical group, and strategic scoring had the highest classification accuracy (78%) (Oetting et al., 2019). In sum, strategic scoring and descriptive evaluation of sentence repetition and sentence production tasks may help identify LI when considering linguistic diversity.

### Considerations in Strategic Scoring

For autistic adolescents and adults, there are considerations in using strategic scoring. One involves evaluating finiteness-marking patterns (Oetting et al., 2016), as findings on finiteness-marking in autism are mixed. While LI in autism includes persistent challenges with structural language, prior work focuses on broad patterns that disallow for focusing on finiteness-marking or adolescents and adults (e.g., Girolamo et al., 2023b). For instance, Modyanova and colleagues (2017) documented group differences in youth ranging from 4 to 16 years. There, ALI (NVIQ 40-112) had lower accuracy than ASD (NVIQ 65-151) on norm-referenced probes for English third-person singular (65.3% < 87.8%) and past-tense (67.8% < 92.8%), as well as higher percent bare stem responses (19.7% > 10.8%), unscorable final responses (19% > 1.9%), and percent responses with wrong tense (30.6% > 0.4%) (Modyanova et al., 2017; Rice & Wexler, 2001). Given this age range, it is unclear whether bare stem responses might be associated with potential dialectal variation or dynamic changes in the language system (Modyanova et al., 2017). A second consideration involves having a sufficient number of productions of morphosyntactic structures to determine finiteness-marking patterns (Oetting et al., 2021). With little precedent and no one standard for finiteness-marking across dialects (Oetting et al., 2019), examining item-level responses and morphosyntactic structures is prudent.

## Summary

Linguistic sensitivity to finiteness-marking in language assessment of autistic individuals is of clinical and scientific relevance. Evidence to date supports the utility of: 1) sentence repetition and sentence production tasks to assess structural language in autism, 2) finiteness-marking to identify LI in autism, and 3) strategic scoring and descriptive examination of utterances and morphosyntactic structures to characterize finiteness-marking. A next step is applying these methods to characterize language in diverse autistic adolescents and adults.

## The Current Study

This study extends the work of Oetting and colleagues (2019; 2016; 2021) to diverse adolescent and adult ALI and ASD ranging in NVIQ. Research questions were:

1. Do scoring method (unmodified and strategic), group (ALI, ASD), and scoring method by group predict item-level scores on sentence repetition and sentence production?
2. Do ALI and ASD groups differ in percent grammatical utterances, percent utterances with zero marking, and percent utterances with wrong tense on sentence repetition and sentence production?
3. Do ALI and ASD groups differ in percent productions of overt, zero, and other responses of: third-person singular–regular and irregular, past-tense–regular and irregular, auxiliary BE, and copula BE on sentence repetition and sentence production?

There were no hypotheses about differences in strategic and unmodified scores, as prior work documenting the clinical utility of strategic scoring was based on significantly younger youth (ages 4-6; Oetting et al., 2016, 2019, 201) and finiteness-marking patterns in autistic youth (ages 4-16) included a wide age range when changes in the language system (including acquisition of finiteness-marking) are dynamic (Modyanova et al., 2017; Rice et al., 1998); thus, disentangling developmental effects from finiteness-marking patterns associated with potential dialectal variation is impossible. Given recent findings from English-speaking ALI that included adolescents with NVIQ < 70 (Modyanova et al., 2017), it was expected that, when considering scoring method, percent grammatical utterances would be lower in the ALI than the ASD group. It was also hypothesized that ALI would have a higher percent of utterances with zero-marking and wrong tense. Finally, it was expected that lower percent overt-marked structures combined and separate would be lower in the ALI than the ASD group.

## Method

This preregistered study (https://osf.io/hzuc4) received institutional board approval. With no precedent of strategic scoring of sentence repetition and sentence production in diverse autistic adolescents and adults, this report examines a subset of participants.

### Selection Criteria

Selection criteria were: (a) ages 13 to 30 years; (b) meet diagnostic criteria for DSM-5 autism (American Psychiatric Association, 2013), per a formal medical or educational diagnosis and independent confirmation using expert clinical judgment plus assessment; (c) use primarily spoken language to communicate, as determined during screening, since study tasks required use of spoken language; and (d) proficiency in American English per self-report during screening, as study tasks were conducted in American English. Participants who did not have sufficient hearing or vision thresholds for hearing and seeing audiovisual stimuli were excluded, as tasks used audiovisual stimuli. Per prior work (Girolamo & Rice, 2022; Girolamo et al., 2023b; Tomblin et al., 1997), the cutoff for ALI was −1.25 *SD* on CELF-5 core language (Wiig et al., 2013) or −1.25 *SD* on at least two measures of overall expressive language, overall receptive language, expressive vocabulary, receptive vocabulary, and nonword repetition (see *Measures*).

### Procedures

Participants were recruited and assessed through two larger studies from 2021 to 2022 (Eigsti & Fein, 2018; Girolamo et al., 2023a). Participants received compensation for their time and effort. Trained examiners administered direct behavioral assessments to participants remotely following test developer guidance (Pearson, 2023). After assessment, CELF-5 (Wiig et al., 2013) Formulated Sentences and Recalling Sentences responses were transcribed, coded, and scored in SALT 20 (Miller & Iglesias, 2020). A trained research assistant ignorant of the study purpose or participant clinical status independently checked point-by-point accuracy. Training involved establishing reliability in a multi-step process: training on transcription and coding manuals; reaching 85% reliability on transcription for utterances and words on three consecutive transcripts; reaching 90% reliability on codes and morphemes on three consecutive transcripts; and reaching 90% reliability on scoring of three consecutive transcripts. Monitoring reliability took place on 20% of data randomly selected for checks by a third, independent trained research assistant; the assumption was that if an examiner did not meet these ongoing checks, they would re-train until they re-established reliability. In this case, research assistants met ongoing reliability checks. All disagreements were discussed until consensus was reached. This procedure resulted in 100% interrater reliability for utterances (as by nature of these tasks, utterances were typically one sentence), 98% for words, and 97.83% for codes and morphemes.

### Measures

Characterizing measures included participant demographics: chronological age, race, ethnicity, sex assigned at birth, and gender. Other measures characterized individual differences: CELF-5 core language score for overall expressive-receptive language ability (Wiig et al., 2013), ADOS-2 calibrated severity scores (Lord et al., 2012) or SRS-2 total *t-*scores (Constantino, 2012) for autism traits, and Wechsler Abbreviated Scale of Intelligence full-scale IQ (Wechsler, 1999) or Raven’s 2 Progressive Matrices–Clinical Edition NVIQ (Raven et al., 2018) for cognitive ability. LI status was determined using a cutoff of −1.25 *SD* on CELF-5 core language (Wiig et al., 2013) or −1.25 *SD* on at least two measures: CELF-5 Expressive Language Index, CELF-5 Receptive Language Index (Wiig et al., 2013), Expressive Vocabulary Test-3^rd^ Ed. (Williams, 2019), Peabody Picture Vocabulary Test-5^th^ Ed. (Dunn, 2019), and the Syllable Repetition Task (Shriberg et al., 2009). Characterizing information did not include language background, as the aim was to evaluate finiteness-marking and not to test differences by dialect.

Measures in analysis came from item responses from the CELF-5 Formulated Sentences and Recalling Sentences (Wiig et al., 2013). For the first research question, the primary outcome was the effect of scoring method (strategic versus unmodified) on item-level score differences. In the second research question, primary outcomes were group differences in percent grammatical utterances, percent zero-marked utterances, and percent utterances with wrong tense. Primary outcomes of the third research question were group differences in percent production of overt, zero, and other structures for third-person singular, past-tense, copula BE, and auxiliary BE.

### Data Processing

#### Coding

Responses were coded for overt and omission of overt finiteness-marking in the following morphosyntactic structures per SALT conventions (Miller & Iglesias, 2020): third person singular regular and irregular, past-tense regular and irregular, auxiliary BE, and copulas (Oetting et al., 2019; Oetting et al., 2016; Oetting et al., 2021). Here, there were only final responses (versus multiple attempts at an item-level response), and coding for omission of overt marking was used to evaluate finiteness-marking patterns versus accuracy. Responses were also coded using the SALT (Miller & Iglesias, 2020) error code for excluded (i.e., other) responses per the schema of Oetting et al. (2021), such as no verb (e.g., “boy”), no subject (e.g., “eats”), wrong tense (e.g., “was eating” or “were eating” for a past-tense form), or some other response (e.g., no response or “I don’t know”). Coded transcripts were used to generate frequencies of production for each morphosyntactic structure (overt, zero or omission of overt-marking, other), as well as utterance types (grammatical, omission of overt marking, wrong tense). Error codes were manually inspected for use of wrong tense or other response.

#### Scoring

Using the CELF-5 manual (Wiig et al., 2013), Formulated Sentences and Recalling Sentences items received unmodified and strategic scores of 0 to 2 or 0 to 3, respectively. Scoring followed manual instructions in terms of number of deviations from the target response. Formulated Sentences responses received scores of 2 if they were complete sentences, semantically, syntactically, and pragmatically appropriate, and included the exact stimulus word; scores of 1 if they met all criteria except for one or two deviations in syntax or semantics; and scores of 0 if responses did not include the stimulus word, were incomplete or illogical sentences, were completely unrelated to the stimulus picture, or had three or more semantic or syntactic deviations (Wiig et al., 2013). Recalling Sentences responses received scores of 3 if sentences had no deviations from the stimulus sentence; scores of 2 if there was one deviation in terms of word changes, additions, substitutions, omissions, or transpositions; scores of if there were 2 or 3 deviations; and scores of 0 if there were four or more deviations (Wiig et al., 2013).

The difference in scoring method involved what was considered a deviation. Again, unmodified scoring only considers GAE norms for finiteness-marking (Oetting et al., 2019). In contrast, strategic scoring accounted for dialectal variation in finiteness-marking that does not confound identification of LI (Oetting et al., 2016). In response to (2a), unmodified scoring counts “work” as a deviation, as in GAE, third-person singular forms must have overt finiteness-marking (i.e., “works”); see (2b). Together with deviations from “nurse,” “community,” and “clinic,” the score would be zero. As in (2c), strategic scoring would not consider “workØ” as a deviation, as omission of overt marking (or zero marking) is acceptable in some dialects, and result in a score of one. After scoring each item-level response, item-level unmodified and strategic scores were translated into overall scores for each subtest.

(2) Example of unmodified and strategic scores from the CELF-5 (Wiig et al., 2013)
(2a) my mother is the nurse who works in the community clinic [stimulus]
(2b) my mother is the <u>woman</u> who <u>work</u> in the <u>place</u> [unmodified] = 4 deviations, score of 0
(2c) my mother is the <u>woman</u> who workØ in the <u>place</u> [strategic] = 3 deviations, score of 1

### Analyses

All responses were included, and basal scores were imputed following CELF-5 scoring rules (Wiig et al., 2013), which mimics real-world practice. One ASD participant had missing information on race and full-scale IQ. No other data were missing. Prior to analysis, variables were inspected to see that they met assumptions of normality, linearity, and heteroscedasticity. Data that did not meet these assumptions used nonparametric analysis. All analyses used an *a priori* significance level of *p* < .05. To address the first research question, scoring condition (unmodified, strategic), group (ASD, ALI) and group by scoring condition were regressed on item-level strategic and unmodified scores of each subtest: CELF-5 Formulated Sentences and Recalling Sentences. This analysis tested for an effect of scoring condition on item-level outcomes and whether groups differed in the effect of condition. To address the second research question, Welch independent sample *t*-tests analyzed group differences in percent grammatical utterances, percent utterances with omission of overt marking, and percent utterances with wrong tense; in effect, utterances were item-level responses. To address the third research question, Welch independent sample *t*-tests analyzed group differences in percent production of overt structures, zero structures, and other responses for third-person singular regular and irregular, past-tense regular and irregular, auxiliary BE, copula BE, and auxiliary DO.

## Results

### Participants

Participants were 31 autistic adolescents and adults (*M*_age_ = 20.80, *SD* = 4.28, 14-30 years); see Table 1. ALI (*n*=15) and ASD (*n*=16) groups did not significantly differ in age. In the full sample, 61.3% of participants were racially minoritized per U.S. Census categories (Office of Management and Budget, 1997): 3.2% Asian, 41.9% Black, 9.7% multiracial, 6.5% Native American, and 29% white. In the ALI group, two participants selected “don’t know” for race and reported they were Puerto Rican. About one quarter (22.6%) of participants were Hispanic or Latine. Most participants were male for sex assigned at birth (female: 19.4%, male: 80.6%), which is similar to male-to-female estimates in autism of 3:1 to 4:1 (Loomes et al., 2017), and gender (female: 22.6%, male: 77.4%). Due to small sample size, Fisher’s exact test was used and revealed no significant group differences in sex assigned at birth, *p* = .172, or gender, *p* = .083.

**Table 1:**
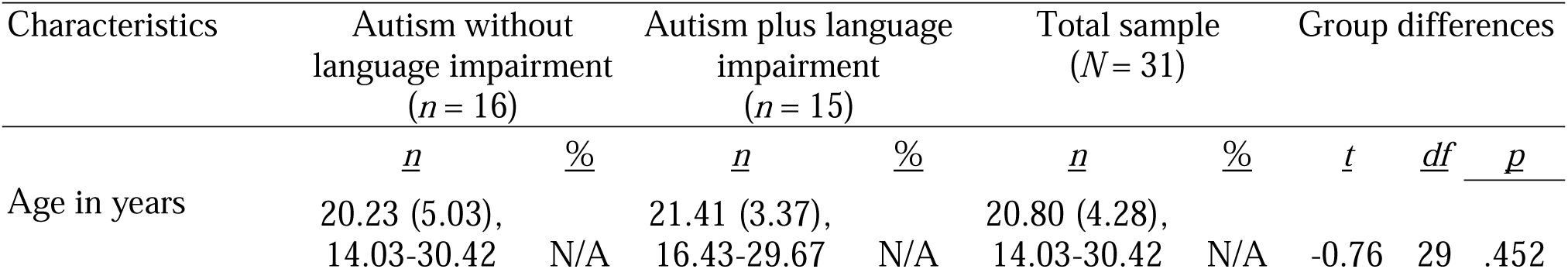
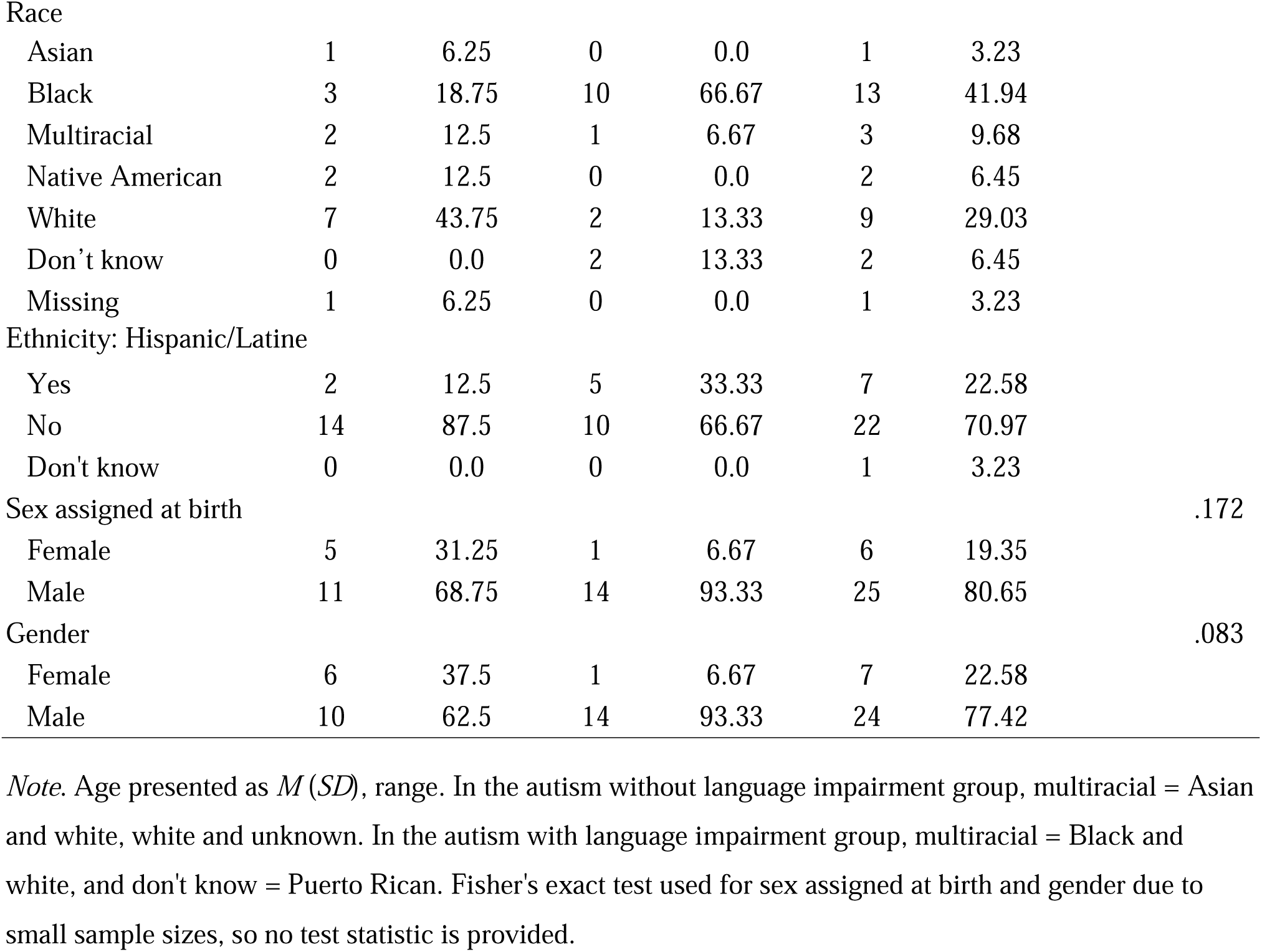
Participant Sociodemographics.

Per grouping criteria, language scores differed; see Table 2. ALI had significantly lower outcomes than the ASD group on CELF-5 core language standard scores (56.2 versus 101.4), Formulated Sentences scaled scores (3.4 versus 11), and Recalling Sentences scaled scores (2.53 versus 9.92). ADOS-2 calibrated severity scores met cutoffs for ASD in both groups (Lord et al., 2012). Mean SRS-2 scores corresponded to “severe” autistic traits in the ASD group and “moderate” autistic traits in the ALI group (Constantino, 2012). For participants with available IQ in the ASD group (*n* = 15), the lowest full scale IQ or NVIQ was 80; NVIQ was −1 to −2 *SD* for two participants (12.5%). In the ALI group, NVIQ was < 70 for three participants (20%) and −1 to −2 *SD* for three participants (20%).

**Table 2:** Participant Language and Nonverbal Cognitive Characteristics.

|  | Autism without<br>language impairment<br>(*n* = 16) |  |  | Autism plus language<br>impairment<br>(*n* = 15) |  |  | Group differences |  |  |
| --- | --- | --- | --- | --- | --- | --- | --- | --- | --- |
|  | *M* | *SD* | *range* | *M* | *SD* | *range* | *t* | *df* | *p* |
| CELF-5 Core Language Score | **101.4** | **13.2** | **82-130** | **56.2** | **12.89** | **40-78** | **8.96** | **25** | **<.001** |
| CELF-5 Formulated Sentences | **11** | **2.22** | **7-16** | **3.4** | **3.54** | **1-11** | **6.48** | **25** | **<.001** |
| CELF-5 Recalling Sentences | **9.92** | **2.71** | **5-15** | **2.53** | **1.92** | **1-6** | **8.28** | **25** | **<.001** |
| Autism traits |  |  |  |  |  |  |  |  |  |
| ADOS-2 calibrated severity score | 7.6 | 1.96 | 4-10 | 8 | N/A | N/A |  |  | N/A |
| SRS-2 total *t*-score | 80.25 | 12.5 | 64-90 | 68.2 | 9.14 | 54-87 |  |  | N/A |
| Cognitive ability |  |  |  |  |  |  |  |  |  |
| Full-scale IQ | 121.1 | 18.4 | 84-152 | 84 | N/A | N/A |  |  | N/A |
| Raven's 2 NVIQ | 92 | 11 | 80-106 | 81.4 | 15.3 | 52-101 |  |  | N/A |

*Note.* Significant differences at $p < .05$ in bolded text. ADOS-2 scores (Lord et al., 2012) reported for 11 participants: 10 autism without language impairment (ASD) and one autism plus language impairment (ALI). Social Responsiveness Scale-2nd Ed. (Constantino et al., 2012) total *t*-scores reported for 17 participants: 4 ASD and 13 ALI. Full scale IQ assessed using the Wechsler Abbreviate Scale of Intelligence (Wechsler, 1999) for 10 participants: 9 ASD and 1 ALI. Nonverbal IQ assessed using the Raven's 2 Progressive Matrices-Clinical Edition (Raven et al., 2018) for 20 participants: 3 ASD and 15 ALI. N/A = not applicable. Independent samples *t*-test used for CELF-5 core language score, Formulated Sentences, and Recalling Sentences.

### Effect of Scoring Method, Group, and Scoring Method by Group on Item-Level Scores

To address the first research question, analyses explored effects of scoring method (unmodified and strategic), group (ALI and ASD), and the interaction of scoring method by group on item-level scores on CELF-5 Formulated Sentences and Recalling Sentences. There was no effect of scoring method, with essentially constant item-level unmodified scores and strategic scores; see Table 3. However, there was a significant effect of group. Welch’s two-sample *t*-tests showed mean item-level scores in the ALI group were significantly lower than the ASD group: Formulated Sentences (0.96 < 1.67) and Recalling Sentences (0.52 < 1.99). Thus, item-level scores differed on the basis of group but not differences in finiteness-marking that could potentially indicate dialectal variation.

**Table 3:** Group Differences in CELF-5 Formulated Sentences and Recalling Sentences Item-Level Strategic and Unmodified Scores, Percent Grammatical Utterances, Percent Omission, and Percent Wrong Tense.

|  | Autism without<br>language<br>impairment<br>(*n* = 16) |  | Autism plus<br>language<br>impairment<br>(*n* = 15) |  | Group differences |  |  |
| --- | --- | --- | --- | --- | --- | --- | --- |
|  | *M* | *SD* | *M* | *SD* | *t* | *df* | *p* |
| CELF-5 Formulated Sentences |  |  |  |  |  |  |  |
| number of utterances | 14.5 | 3.83 | 12.8 | 7.13 | 0.83 | 29 | .411 |
| unmodified score | **1.67** | **0.19** | **0.96** | **0.49** | **5.23** | **18.23** | **< .0001** |
| strategic scores | **1.67** | **0.18** | **0.96** | **0.49** | **5.3** | **17.84** | **< .0001** |
| Percent Grammatical Utterances | **94.95** | **8.42** | **77.93** | **23.5** | **2.57** | **15.91** | **.021** |
| Omission | 1.26 | 3.56 | 9.23 | 20.16 | -1.46 | 13.71 | .167 |
| Wrong Tense | 0.97 | 2.65 | 1.77 | 4.65 | -0.59 | 28 | .560 |
| CELF-5 Recalling Sentences |  |  |  |  |  |  |  |
| number of utterances | 15 | 4.9 | 16.8 | 6.86 | -0.85 | 29 | .405 |
| unmodified score | **1.99** | **0.46** | **1.32** | **0.52** | **3.73** | **27.6** | **.001** |
| strategic scores | **1.99** | **0.46** | **1.32** | **0.52** | **3.73** | **27.6** | **.001** |
| Percent Grammatical Utterances | 92.42 | 7.84 | 79.62 | 22 | 2.07 | 15.88 | .056 |
| Omission | 3.11 | 4.35 | 3.02 | 5.23 | 0.05 | 28 | .963 |
| Wrong Tense | 1.14 | 2.51 | 2.35 | 4.71 | -0.86 | 19.23 | .401 |

*Note.* Significant differences at $p < .05$ in bolded text. Omission = omission of overt finiteness-marking. Welch's two sample *t*-test used for unmodified and strategic scores on Formulated Sentences and Recalling Sentences, percent grammatical utterances on both subtests Formulated Sentences and Recalling Sentences, omission on CELF-5 Formulated Sentences, and wrong tense on CELF-5 Recalling Sentences. Percentages do not add up to 100%, as some responses were "other."

Next, regression models were run to predict item-level Formulated Sentences and Recalling Sentences scores from group, scoring method, and an interaction of group by scoring method. Each multiple regression model significantly predicted outcomes, Formulated Sentences scores, *F*(3,50) = 17.17, *p* < .0001, and Recalling Sentences scores, *F*(3,50) = 9.51, *p* < .00. In addition, there were main effects of group (*p*’s < .0001), but effects of scoring method and group by scoring method were non-significant; see Table 4. Findings indicate that scores differed by LI status but not by scoring method. Thus, differences in finiteness-marking associated with dialectal variation did not confound outcomes.

**Table 4.** Regression Results for CELF-5 Formulated Sentences and Recalling Sentences Strategic Versus Unmodified Scores.

| Variable | Model 1 |  |  |  | Model 2 |  |  |  |
| --- | --- | --- | --- | --- | --- | --- | --- | --- |
|  | *B* | *SE* | *t* | *p* | *B* | *SE* | *t* | *p* |
| CELF-5 Formulated Sentences |  |  |  |  |  |  |  |  |
| Constant | 1.30 | 0.10 | 12.74 | < .0001 | 1.71 | 0.11 | 15.67 | < .0001 |
| Unmodified vs. strategic score | -0.003 | 0.14 | -0.02 | .983 | -0.01 | 0.15 | -0.05 | .964 |
| Group |  |  |  |  | -0.75 | 0.15 | -5.10 | < .0001 |
| Group by scoring condition |  |  |  |  | 0.01 | 0.21 | 0.03 | .973 |
| Adjusted $R^2$ | -0.02 | | | | 0.48 | | | < .0001 |
| CELF-5 Recalling Sentences |  |  |  |  |  |  |  |  |
| Constant | 0 | 0 | 14.05 | < .0001 | 0 | 0 | 14.33 | < .0001 |
| Unmodified vs. strategic score | 0 | 0 | 0 | 1 | 0 | 0 | 0 | 1 |
| Group |  |  |  |  | 0 | 0 | -3.78 | < .0001 |
| Group by scoring condition |  |  |  |  | 0 | 0 | 0 | 1 |
| Adjusted $R^2$ | -0.02 | | | | 0.33 | | | < .0001 |

*Note.* Group = autism without language impairment or autism plus language impairment. Scoring condition = unmodified scoring or strategic scoring.

### Group Differences in Percent Grammatical Utterances, Percent Utterances with Overt Omission, and Percent Utterances with Wrong Tense in Sentence Repetition and Sentence Production

To contextualize responses, analyses for the second research question examined group differences in percent grammatical utterances, percent utterances with omission of overt marking, and percent utterances with wrong tense. ALI and ASD groups did not significantly differ in number of utterances on Formulated Sentences (12.8 versus 14.5) or Recalling Sentences (16.8 versus 15); see Table 3.

Groups significantly differed in percent grammatical utterances on Formulated Sentences (77.93% versus 94.95%); see Figure 1 and Table 3. However, ALI and ASD groups did not significantly differ on percent utterances with omission of overt marking or with use of wrong tense: Formulated Sentences wrong tense (1.77% versus 0.97%), Recalling Sentences omission of overt marking (3.02% versus 3.11%), and Recalling Sentences wrong tense (2.35% versus 1.14%). Other group differences were not significant: Formulated Sentences percent utterances with omission of overt marking (ALI: 9.23% versus ASD: 1.26%) and Recalling Sentences percent grammatical utterances (ALI: 79.62% versus ASD: 92.42%). In sum, the ALI group had significantly lower percent grammatical utterances on sentence production than the ASD group but not on other measures.

**Figure 1.**
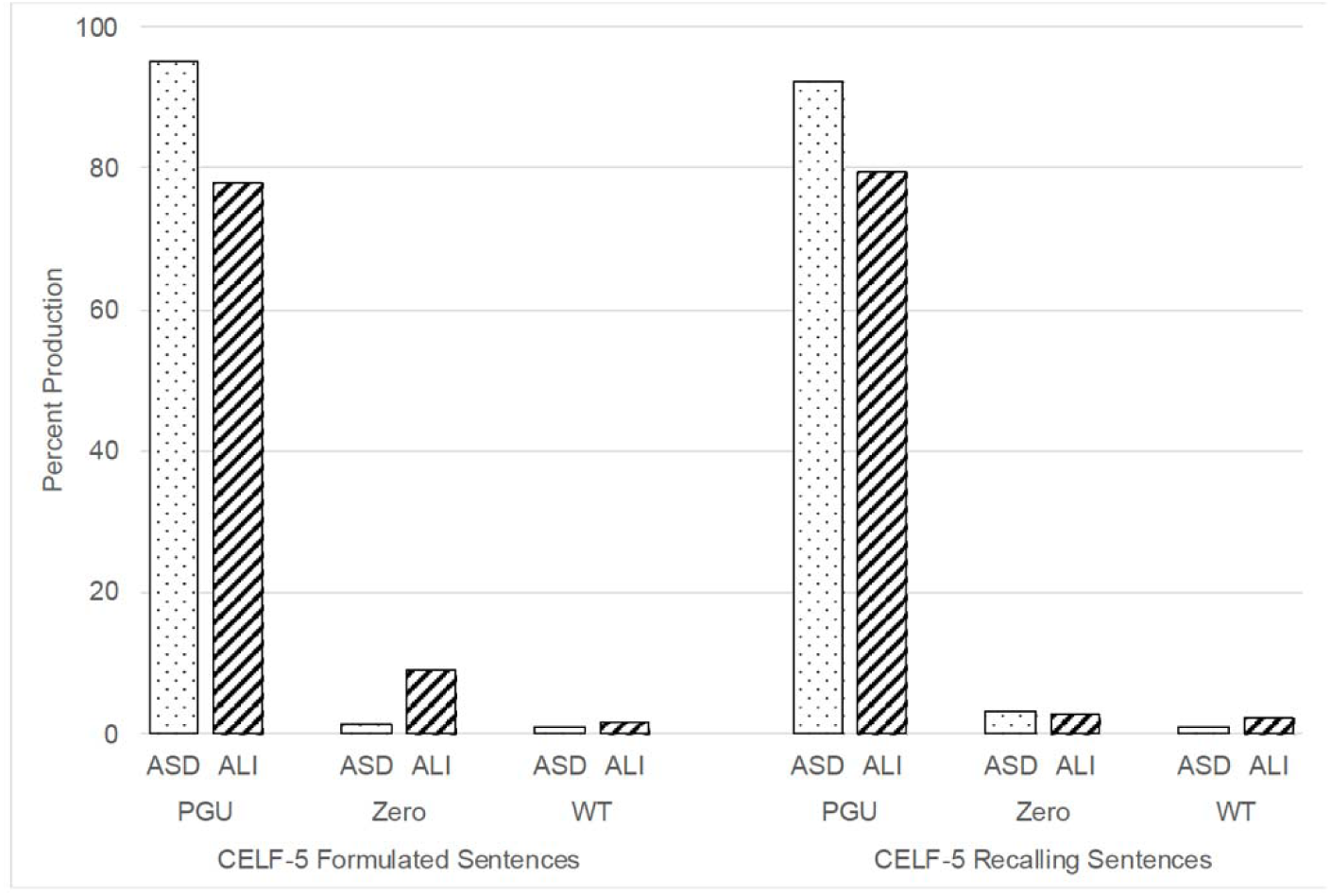
Percent grammatical utterances, percent utterances with omission of overt finiteness marking, and percent utterance with wrong tense. Note. ASD = autism without language impairment. ALI = autism plus language impairment. PGU = percent grammar

### Group Differences in Percent Productions of Overt, Zero, and Other Responses of Finiteness-Marking Structures

The third research question tested group differences in percent production of overt, zero, and other responses of finiteness-marking patterns of morphosyntactic structures combined and separately that differentiate LI across dialects of American English (Oetting et al., 2016, 2019, 2021). Specific structures were: third-person singular regulars and irregulars, past-tense regulars and irregulars, auxiliary BE, and copulas.

#### Finiteness-Marking in Morphosyntactic Structures Combined

When considering structures together, ALI and ASD groups did not significantly differ in number of morphosyntactic structures produced on CELF-5 Formulated Sentences (15.13 versus 15), or CELF-5 Recalling Sentences (18.93 versus 22.92); see Table 5. ALI and ASD groups both produced structures with overt marking >90% on Formulated Sentences (92.69% versus 99.4%) and Recalling Sentences (90.06% versus 98.8%); see Figure 2. However, the ALI group had significantly lower percent structures with overt marking than the ASD group on Recalling Sentences. Groups did not significantly differ on other outcomes. On both Formulated Sentences and Sentence Repetition, percent structures with omission of overt marking (ALI: 6.38% and 5.23% versus ASD: 0.6% and 0.4%) and percent structures with other productions on (ALI: 0.93% and 4.71% versus ASD: 0% and 0.78%) were low. Overall, group differences were minimal, with overt marking above 90% on both sentence production and sentence repetition.

**Figure 2.**
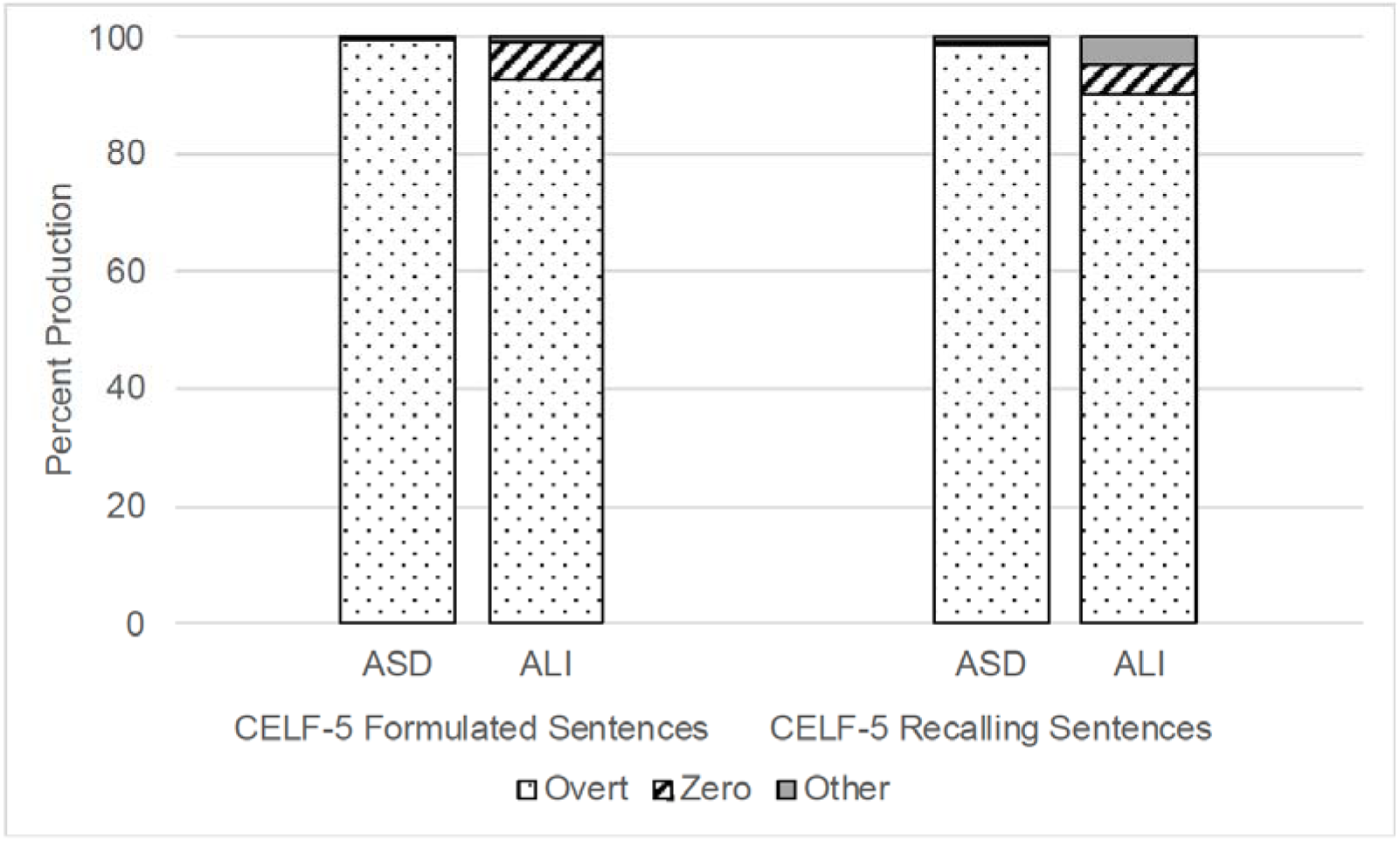
Percent overt marking, zero marking, and other production of morphosyntactic structures combined. Note. ASD = autism without language impairment. ALI = autism plus language impairment. Overt = overt finiteness-marking. Zero = omission of overt marking. Other = other production (e.g., no verb).

**Table 5:**
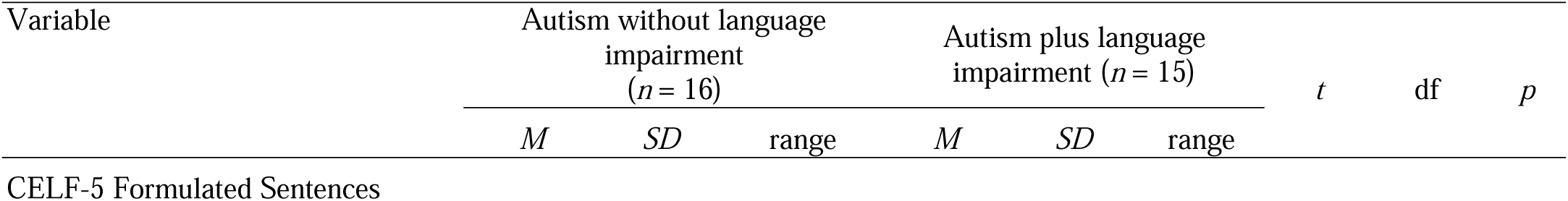
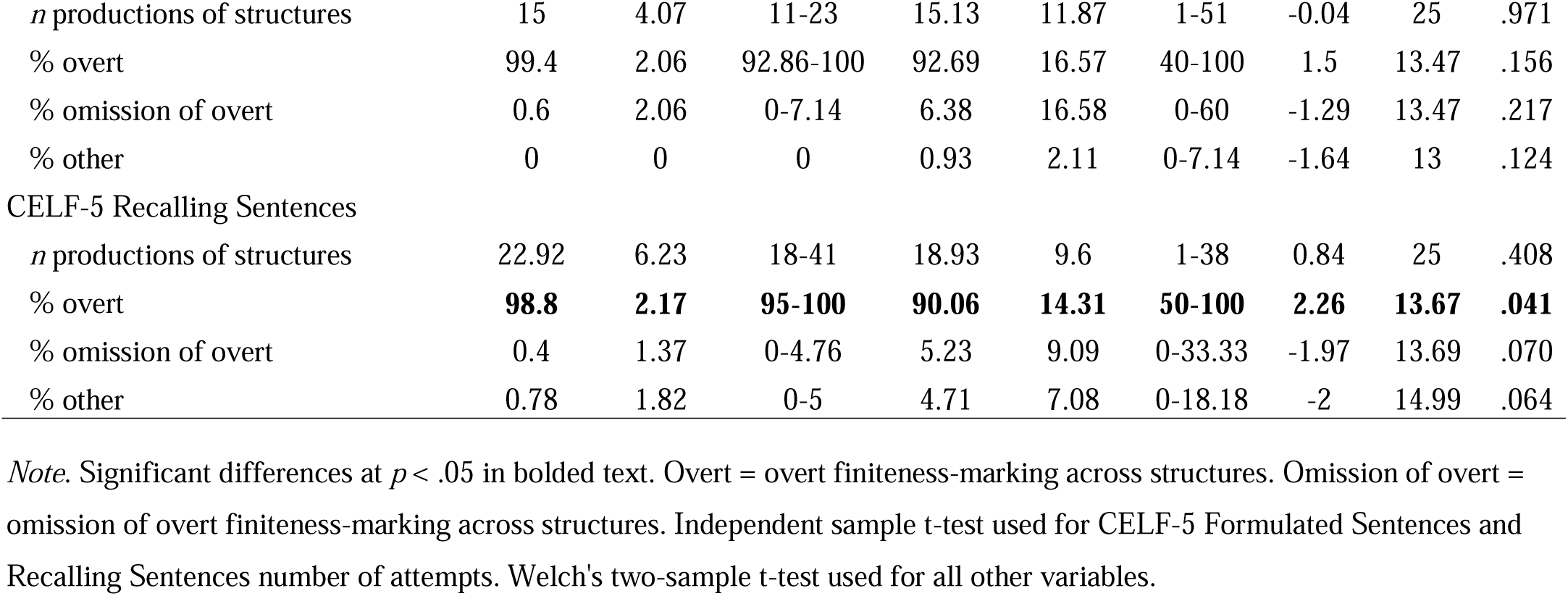
Group Differences in Finiteness-Marking on All Morphosyntactic Structures Combined on CELF-5 Formulated Sentences and Recalling Sentences (Wiig et al., 2013)

| Variable | Autism without language impairment<br>(*n* = 16) |  |  | Autism plus language impairment (*n* = 15) |  |  | *t* | df | *p* |
| --- | --- | --- | --- | --- | --- | --- | --- | --- | --- |
|  | *M* | *SD* | range | *M* | *SD* | range |  |  |  |
| CELF-5 Formulated Sentences |  |  |  |  |  |  |  |  |  |
| *n* productions of structures | 15 | 4.07 | 11-23 | 15.13 | 11.87 | 1-51 | -0.04 | 25 | .971 |
| % overt | 99.4 | 2.06 | 92.86-100 | 92.69 | 16.57 | 40-100 | 1.5 | 13.47 | .156 |
| % omission of overt | 0.6 | 2.06 | 0-7.14 | 6.38 | 16.58 | 0-60 | -1.29 | 13.47 | .217 |
| % other | 0 | 0 | 0 | 0.93 | 2.11 | 0-7.14 | -1.64 | 13 | .124 |
| **CELF-5 Recalling Sentences** |  |  |  |  |  |  |  |  |  |
| *n* productions of structures | 22.92 | 6.23 | 18-41 | 18.93 | 9.6 | 1-38 | 0.84 | 25 | .408 |
| % overt | **98.8** | **2.17** | **95-100** | **90.06** | **14.31** | **50-100** | **2.26** | **13.67** | **.041** |
| % omission of overt | 0.4 | 1.37 | 0-4.76 | 5.23 | 9.09 | 0-33.33 | -1.97 | 13.69 | .070 |
| % other | 0.78 | 1.82 | 0-5 | 4.71 | 7.08 | 0-18.18 | -2 | 14.99 | .064 |

*Note.* Significant differences at $p < .05$ in bolded text. Overt = overt finiteness-marking across structures. Omission of overt = omission of overt finiteness-marking across structures. Independent sample t-test used for CELF-5 Formulated Sentences and Recalling Sentences number of attempts. Welch's two-sample t-test used for all other variables.

#### Finiteness-Marking in Individual Morphosyntactic Structures

As a final check, analyses examined finiteness-marking in individual morphosyntactic structures: third-person singular regulars and irregulars, past-tense regulars and irregulars, auxiliary BE, and copulas. Because frequencies of individual structures were low (e.g., auxiliary *be* singular present; range: 0-3.83), forms were combined into third-person regular and irregular, past-tense regular and irregular, auxiliary BE, and copula BE in order to analyze finiteness-marking patterns (Oetting et al., 2019); see Tables 6 and 7. Groups differed in number of productions for auxiliary BE present on Formulated Sentences (ALI: 3.73 > ASD: 1.25), *t*(25) = −2.54, *p* = .018; auxiliary BE present on Recalling Sentences (ALI: 0.87 > ASD: 0.08), *t*(24) = - 3.44, *p* = .002; and past-tense regular and irregulars (ASD: 16 > ALI: 7.27), *t*(18.32) = 4.85, *p* < .001 on Recalling Sentences.

**Table 6:** Total and Mean Production of Morphosyntactic Structures on CELF-5 Formulated Sentences and Recalling Sentences.

|  | total<br>*N* | Autism without language<br>impairment<br>(*n* = 16) | Autism plus language<br>impairment<br>(*n* = 15) |
| --- | --- | --- | --- |
|  |  | <u>*M*</u> | <u>*M*</u> |
| CELF-5 Formulated Sentences |  |  |  |
| 3s regular | 82 | 3.83 | 2.4 |
| 3s - irregular | 15 | 0.67 | 0.47 |
| Past regular | 47 | 2.42 | 1.2 |
| Past irregular | 67 | 2.33 | 2.6 |
| Auxiliary singular present | 53 | 1.08 | 2.67 |
| Auxiliary plural present | 18 | 0.17 | 0.08 |
| Auxiliary singular past | 8 | 0.33 | 0.27 |
| Auxiliary plural past | 3 | 0.17 | 0.07 |
| Copula singular present BE | 66 | 2.42 | 2.47 |
| Copula plural present BE | 15 | 0.33 | 0.73 |
| Copula singular past BE | 16 | 0.5 | 0.67 |
| Copula plural past BE | 6 | 0.33 | 0.13 |
| CELF-5 Recalling Sentences |  |  |  |
| 3s regular | 44 | 3.83 | 2.4 |
| 3s - irregular | 2 | 0.67 | 0.47 |
| Past regular | 174 | 2.42 | 1.2 |
| Past irregular | 127 | 2.33 | 2.6 |
| Auxiliary singular present | 2 | 1.08 | 2.67 |
| Auxiliary plural present | 12 | 1.07 | 0.73 |
| Auxiliary singular past | 0 | - | - |
| Auxiliary plural past | 0 | - | - |
| Copula singular present BE | 57 | 2.42 | 2.47 |
| Copula plural present BE | 3 | 0.33 | 0.73 |
| Copula singular past BE | 57 | 0.5 | 0.67 |
| Copula plural past BE | 34 | 0.33 | 0.13 |

*Note.* - indicates no mean could be calculated due to no productions of a structure.

**Table 7:** Overt, Zero, and Other Productions of Morphosyntactic Structures by Group.

| Structure | Total Structures |  | Autism without language impairment (n=16) |  |  | Autism plus language impairment (n=15) |  |  |
| --- | --- | --- | --- | --- | --- | --- | --- | --- |
|  | N | M | overt | zero | other | overt | zero | other |
| CELF-5 Formulated Sentences |  |  |  |  |  |  |  |  |
| 3s regular & irregular | 97 | 3.59 | 98.86 | 1.14 | 0 | 81.25 | 18.75 | 0 |
| Past regular & irregular | 114 | 4.22 | 100 | 0 | 0 | 100 | 0 | 0 |
| Auxiliary BE present | 71 | 2.63 | 100 | 0 | 0 | 100 | 0 | 0 |
| Auxiliary BE past | 11 | 0.41 | 100 | 0 | 0 | 83.33 | 0 | 16.67 |
| Copula present | 81 | 3 | 100 | 0 | 0 | 97.08 | 0 | 2.92 |
| Copula past | 22 | 0.81 | 100 | 0 | 0 | 100 | 0 | 0 |
| CELF-5 Recalling Sentences |  |  |  |  |  |  |  |  |
| 3s regular & irregular | 46 | 1.7 | 95.83 | 4.17 | 0 | 81.94 | 18.06 | 0 |
| Past regular & irregular | 301 | 11.15 | 99.54 | 0 | 0.46 | 98.1 | 1.9 | 0 |
| Auxiliary BE present | 14 | 0.52 | - | - | - | 100 | 0 | 0 |
| Auxiliary BE past | 0 | - | - | - | - | - | - | - |
| Copula present | 60 | 2.22 | 100 | 0 | 0 | 100 | 0 | 0 |
| Copula past | 91 | 3.37 | 98.83 | 0 | 1.17 | 75.98 | 3.27 | 20.75 |

*Note.* ALI = autism plus language impairment. ASD = autism without language impairment. *M* not possible, as auxiliary BE present involved.

Analyses showed no group differences in finiteness-marking patterns of third-person regulars and irregulars, past-tense regulars and irregulars, auxiliary BE, or copulas; see Tables 7 and 8. The ALI group had lower percent overt production of third-person (81.25% versus 98.86%) and auxiliary BE past (83.33% versus 100%) on Formulated Sentences, as well as third-person (81.94% versus 95.83%) and past-tense copulas (75.98% versus 98.83%) on Recalling Sentences than the ASD group, but these differences were not significant. Similarly, the ALI group had higher percent production of zero forms than the ASD group on third person on Formulated Sentences (18.75% versus 1.14%) and Recalling Sentences (18.06% versus 4.17%), as well as of other production for past-tense copula on Recalling Sentences (20.75% versus 1.17%). Altogether, groups did not differ in percent productions of overt, omission of overt, or other production of third-person, past tense, auxiliary, or copula structures.

**Table 8:** Statistical Comparisons of Production of Overt, Zero, and Other Morphosyntactic Structures by Group.

| Structure | overt |  |  | zero |  |  | other |  |  |
| --- | --- | --- | --- | --- | --- | --- | --- | --- | --- |
|  | *t* | *df* | *p* | *t* | *df* | *p* | *t* | *df* | *p* |
| CELF-5 Formulated Sentences |  |  |  |  |  |  |  |  |  |
| 3s regular & irregular | 1.57 | 11.23 | .144 | -1.57 | 11.23 | .144 | - | - | - |
| Past regular & irregular | - | - | - | - | - | - | - | - | - |
| Auxiliary BE present | - | - | - | - | - | - | - | - | - |
| Auxiliary BE past | 1 | 2 | .423 | - | - | - | -1 | 2 | .423 |
| Copula present | 1.27 | 10 | .233 | - | - | - | -1.27 | 10 | .233 |
| Copula past | - | - | - | - | - | - | - | - | - |
| CELF-5 Recalling Sentences |  |  |  |  |  |  |  |  |  |
| 3s regular & irregular | -1.39 | 15.46 | .183 | -1.39 | 15.46 | .183 | - | - | - |
| Past regular & irregular | 1.26 | 16.48 | .227 | -1.82 | 12 | .094 | 1 | 11 | .169 |
| Auxiliary BE present | - | - | - | - | - | - | - | - | - |
| Auxiliary BE past | - | - | - | - | - | - | - | - | - |
| Copula present | - | - | - | - | - | - | - | - | - |
| Copula past | 1.77 | 16.97 | .095 | -1.32 | 13 | .209 | -1.53 | 17.26 | .145 |

*Note.* Group = autism without language impairment or autism plus language impairment. 3s = third-person singular present tense. - = t-test not possible. On Formulated Sentences, computing *t* is not possible, because *SDs* of groups are 0. On Recalling Sentences, computing *t* is not possible for auxiliary BE past overt and auxiliary BE past zero, because at least one group is empty. On Recalling Sentences, computing *t* is not possible for copula BE present overt and zero, because the *SDs* of groups are 0.

### Summary

Item-level strategic scores and unmodified scores did not differ on sentence repetition or sentence production. When accounting for scoring method, group, and group by scoring method, only group predicted item-level score outcomes. At the level of utterances and morphosyntactic structures, groups differed in: (a) percent grammatical utterances on sentence production (ALI < ASD) and (b) percent production of overt structures combined on sentence repetition – and percentages were each over 90% (ALI < ASD).

## Discussion

Analyzing sentence production and sentence repetition tasks in diverse autistic adolescents and adults showed no differences in strategic and unmodified scores and limited group differences in utterance-, item- and structure-level outcomes. The motivation was not to prove differences in strategic and unmodified scores, but rather, to understand possible confounds in finiteness-marking. Findings have implications for understanding structural language in autism beyond the school-aged years.

### Comparisons to Published Patterns

In this sample, the ASD group only had significantly higher performance than the ALI group on percent grammatical utterances of sentence production. Percent utterances with omission of overt marking was low in both groups for sentence repetition (∼3%) but qualitatively higher in ALI than ASD for sentence production (9.23% versus 1.26%). One interpretation might be that dialectal variation in finiteness-marking confounded scoring outcomes, as the ALI group was primarily minoritized and the ASD group was primarily white – even though strategic and unmodified scores did not differ. However, recall that group comparisons for each subtest was based on a mean of about 15 utterances (or items). By nature, this number of utterances limits the number of productions of morphosyntactic structures (Oetting et al., 2021). Regardless, the fact that percentage of utterances with omission of overt marking was near 0% indicates that it is *not* the case that minoritized autistic adolescents and adults are universally only proficient in dialects that differ in finiteness-marking norms from GAE (Oetting, 2020).

Results shed light on finiteness-marking in ALI and ASD groups across age ranges. Compared to Modyanova et al. (2017), ALI and ASD groups showed fewer differences. In Modyanova et al. (2017), ALI and ASD groups (ages 6-16) differed in percent total responses (or utterances) with overt finiteness-marking on third-person singular (ALI: 65.3% versus ASD: 87.8%) and past tense (ALI: 67.83% versus ASD: 92.82%) elicitation probes, as well as percent omission on both third-person singular (ALI: 15.1% versus ASD: 7.2%) and past-tense probes (ALI: 19.7% versus ASD: 10.8%) (Modyanova et al., 2017). In this study of older individuals (*M*_age_: 20.80), comparisons were limited by a low number of productions on each task for third-person singular (1.7 to 3.59) and past-tense (4.22 to 11.15). Still, the ALI group had a lower percent production of overt third-person singulars than the ASD group on both sentence production and sentence repetition (81.25% to 81.94% versus 95.83% to 98.86%). Unlike Modyanova et al. (2017), production of overt past-tense structures was near ceiling in both groups on both tasks (range: 98.1% to 100%), while omission of overt marking was low in both utterances (range: 1.26% to 9.23%) and structures (range: 1.37% to 6.38%). While deeper conclusions about finiteness-marking cannot be made without a higher number of productions of utterances or structures, one possibility is that early acquired morphosyntactic forms may be less clinically useful for identifying LI in older autistic individuals (e.g., Rice & Wexler, 2001).

In contrast, our findings do not replicate Manenti et al. (2023), who administered a French sentence repetition task to 39 autistic adults (ages 18-56), one-third of whom had intellectual disability. There, the “ASD-low language” group (*n*=10) had lower finiteness-marking than the “ASD-normal language” group (*n*= 24) (26.7% versus 98.6%) (Manenti et al., 2023). In this study, group differences were much more modest, with overt finiteness-marking in sentence repetition in both groups near ceiling (90.06% versus 98.8%). Differences between studies might explain these differences in outcomes. First, finite clauses in the French sentence repetition task focused on present-tense, whereas the sentence repetition task of this study also included past-tense, auxiliaries, and copulas. A second possible factor involves participant characteristics. Manenti et al. (2023) recruited autistic adults from residential facilities, reporting that six of 10 “ASD-low” participants did not attend “regular school” and that nine of 10 had NVIQ < 68. As only age equivalencies for NVIQ were reported, the variability of NVIQ in the ASD-low sample is unknown. Here, NVIQ was variable in both ALI (52-101) and ASD (80-152) groups, and participants were recruited from the community versus more restrictive settings. Third, as with Modyanova et al. (2017), Manenti et al. (2023) used an experimental probe designed to elicit certain morphosyntactic forms; this study did not. Last, it may be that while finiteness-marking norms in English are well understood in childhood (Rice et al., 1998), norms in adolescence and adulthood differ.

Overall, findings in this study showed that sentence production and sentence repetition outcomes were sensitive to heterogeneity in autistic adolescents and adults varying in NVIQ and language skills. However, as some participants were beyond the age range for the CELF-5 (Wiig et al., 2013), the clinical utility is unknown. One issue is that norm-referenced language assessments designed for adults do not comprehensively measure language, as they are designed to measure the severity of specific acquired disorders, such as the Western Aphasia Battery and the Montreal Cognitive Assessment (Kertesz, 2022; Nasreddine et al., 2005). Further, some assessments are succinct by design to maximize utility for point-of-service clinical design-making in inpatient settings. As such, despite excitement about increasing accessibility of language assessment across the autism spectrum (Kover & Abbeduto, 2023; Schaeffer et al., 2023), best practices for language assessment that is developmentally appropriate and age appropriate in autistic adolescents and adults remains elusive (Howlin & Taylor, 2015).

### Implications for Clinical Practice and Research

Beyond empirical findings, the approach of this study has implications for assessment in clinical and research settings. Clinicians often use norm-referenced assessments, follow workplace eligibility policy, and report limitations in ability to conduct linguistically sensitive assessment (Denman et al., 2021; Selin et al., 2022). One challenge is that assessments sensitive to language disorders across dialects in English, such as the Diagnostic Evaluation of Language Variation (Seymour et al., 2018), only extend to ages 9;11. Though imperfect, for assessments that extend to adolescence and early adulthood, examiners can implement a multi-pronged approach to enhance linguistic sensitivity: evaluation of finiteness-marking patterns (Oetting & Garrity, 2006; Oetting & McDonald, 2001), evaluation of finiteness-marking at the item level (Oetting et al., 2016), and comparison of whether accounting for dialectal variation in finiteness-marking impacts scores on clinical assessment (Oetting et al., 2019). Such an approach can help provide incremental evidence to support clinical decision-making.

In research, methodologies that yield replicable results not only provide data on the soundness of measurement, but also have the potential to support researchers in reducing biases toward groups who are systematically excluded from research (Maye et al., 2022; Russell et al., 2019). Indeed, researchers have the responsibility to ensure research methodologies are accessible and responsive to participants, including linguistic sensitivity (Oetting, 2020). When researchers fail to include feasible methods, such as strategic scoring, they risk perpetuating assumptions often made about the language backgrounds of individuals, solely based on perceptions about individuals or groups (Dunham et al., 2015; Marques et al., 1988, 1998). This responsibility is even more pronounced when considering the overlap between pragmatic language, an area of particular interest in autism research, and structural language (Schaeffer et al., 2023). To combat such bias, researchers can – and should – integrate linguistically sensitive methodological tools into tasks, such as sentence repetition, which provide useful information on structural language in autism-appropriate tasks (Schaeffer et al., 2023).

### Limitations

Although this study focused on a new approach to characterizing finiteness-marking in autism, it had several limitations. First, though unmodified and strategic scores did not differ, participants also produced few morphosyntactic structures that differentiate LI across dialects. Per Oetting et al. (2019), an insufficient frequency distribution disallows from examining patterns that could inform development of probes. In addition, analyzing data from adolescents and adults, when changes in the language system are less dynamic (Howlin, 1984; Miniscalco & Carlsson, 2022), may mask differences in the sensitivity of scoring approaches to dialectal variation and structural language skills in autism. To better understand the clinical utility of strategic scoring, replication with a larger, more developmentally diverse sample is needed.

Second, this study did not include comprehensive information on language background. The purpose was to analyze finiteness-marking using through a multi-step approach, which is congruent with arguments for valid test interpretation and use (Messick, 1990). Per the American Speech-Language-Hearing Association Code of Ethics (2023) and scope of practice in speech-language pathology (2016), examiners may not always have the full range of knowledge about an individual’s language experiences or environment yet are nevertheless responsible for working to linguistically sensitive in assessment. While more robust examination of language variation is needed for language in autism (e.g., Oetting et al., 2016, 2019, 2021), this initial investigation describes one strategy to evaluate patterns and check for potential differences in finiteness-marking associated with dialectal variation that could confound scoring of sentence production or sentence repetition tasks (Oetting & McDonald, 2001). Finally, this study did not examine effects of vocabulary and memory on sentence repetition (Manenti et al., 2023). While relevant to understanding overall sentence repetition performance, this study focused on potential dialectal variation in finiteness-marking.

### Future Directions

This study lays groundwork for future research on structural language in older autistic individuals that is high-quality (Howlin & Taylor, 2015), linguistically sensitive (Oetting, 2020), and inclusive of individuals varying in IQ (Manenti et al., 2023). A broader understanding of finiteness-marking on sentence repetition and sentence production is needed to establish the clinical utility of strategic scoring for diverse autistic individuals (Oetting & McDonald, 2001). Evaluating finiteness-marking in large samples of autistic youth and adults who are chronologically and developmentally diverse would allow for observation of finiteness-marking when changes in the language system are more dynamic (Kwok et al., 2015). This work, which is underway, is relevant in understanding the potential of sentence repetition to sensitively characterize structural language in autism (Schaeffer et al., 2023). An additional future direction for research involves comparison of sentence repetition and sentence production to dialect-sensitive probes that elicit a sufficient number of morphosyntactic structures sensitive to structural language skills across dialects for autistic adolescents and adults (Oetting et al., 2016, 2019, 2021). Such comparison would inform to what extent these tasks, which are common in clinical settings and neuroimaging (Betz et al., 2013; Larson et al., 2023), are clinically useful. These are two of many directions that support achieving a dimensional understanding of language in autism (Kover & Abbeduto, 2023).

## Conclusion

In evaluating finiteness-marking on sentence repetition and sentence production data from diverse autistic adolescents and adults in English, this report offers one way to infuse linguistically sensitivity in common language assessment measures. The aim was not to “prove” differences existed. The take-home point is that carefully evaluating scores, utterance types, and productions of morphosyntactic structures are feasible and ought to inform interpretation and use of assessment data to characterize structural language as a dimensional construct (Kover & Abbeduto, 2023). In the long-term, these efforts may contribute to much-needed reliable and generalizable approaches to language assessment in autistic adolescents and adults.

## Acknowledgements

TG was supported by T32DC017703 (PD: Eigsti), L70DC021323 (PI: Girolamo) & an ASHFoundation New Investigators Research Grant (PI: Girolamo). SG was supported by an ASHFoundation Graduate Student Scholarship & a University of Kansas Graduate Fellowship (PI: Ghali). CL was supported by R01MH112687 (PI: Eigsti). We also thank community partners, participants and their families, research assistants, and lab personnel.

## Data Availability Statement

The datasets generated during the current study are not publicly available due to participants opting not to share their de-identified individual data in the consent process or due to ongoing analyses. Information about the datasets (structure, code) are available from the corresponding author on reasonable request.

## CRediT statement

TG: conceptualization, methodology, formal analysis, investigation, resources, data curation, writing – original draft, project administration, funding acquisition. SG: formal analysis, validation, writing – review & editing. CL: formal analysis, data curation, writing – review & editing, visualization.

## Author Information

Teresa Girolamo, School of Speech, Language, and Hearing Sciences, San Diego, CA, San Diego State University. Samantha Ghali, Child Language Doctoral Program, Lawrence, KS, University of Kansas. Caroline Larson, Department of Speech, Language, and Hearing Sciences, Interdisciplinary Neuroscience Program, Columbia, MO, University of Missouri.

## Funding

TG was supported by T32DC017703 (PD: Eigsti), L70DC021323 (PI: Girolamo) & an ASHFoundation New Investigators Research Grant (PI: Girolamo). SG was supported by an ASHFoundation Graduate Student Scholarship & a University of Kansas Diversity, Equity, Inclusion, and Belonging Graduate Fellowship (PI: Ghali). CL was supported by R01MH112687 (PI: Eigsti).

## Conflict of Interest

The authors have no financial or nonfinancial disclosures.

## Notes

### Competing Interest Statement

The authors have declared no competing interest.

### Author Declarations

IRB of the University of Connecticut gave ethical approval for this work.

