## Supplementary Materials 1 for "Sentence production and sentence repetition in autistic adolescents and young adults: Linguistic sensitivity to finiteness-marking"

| **Table 1**  *Cross-linguistic autism studies published in English employing sentence repetition and sentence production tasks* | | | | | | | | | | | | | | |
| --- | --- | --- | --- | --- | --- | --- | --- | --- | --- | --- | --- | --- | --- | --- |
| Study | Race/Ethnicity | Language | *N* | Dx | ASD age | LI Definition | | ASD NVIQ | Task | | | Group Comparisons | *p* | *g* |
|  |  |  |  |  |  | NVIQ | Language |  | Measure | SR | SP |  |  |  |
| **ASD < TD** | | | | | | | | | | | | | |  |
| Alhassan & Marinis (2021) | NR | Arabic | 10 ASD  42 TD | NR | 5.83 | - | Arabic PPVT ≤ -1 *SD* | 14.6 (4.89) | Saudi-SRT % | x |  | **.23 (.1) < .36 (.2)** | **.012** | **0.62** |
| Arutiunian et al. (2022) | NR | Russian | 60 ASD 25 TD | ICD-10 | 9.11 | - | - | 83.1 (20.5) | KORABLIK % | x |  | **0.7 (0.4) < 1.0 (.1)** | **<.001** | **0.9** |
|  |  |  |  |  |  |  |  |  |  |  | x | **0.6 (0.4) < 1.0 (0.1)** | **<.001** | **1.02** |
| Brynskov et al. (2017) | NR | Danish | 21 ASD 21 TD | ICD-10 | 4-6 | - | - | 106 (19) | CELF-P2 raw score | x |  | **21.4 (11.0) < 29.8 (4.2)** | **.001** | **0.99** |
| Landa & Goldberg (2005) |  | English | 19 "HFA" 19 TD | NR | 11.01 | ≥80 | - | 104.6 (13.5) | CELF-R SS |  | x | **7.2 (2.7) < 9.8 (3.4)** | **<.01** | **0.82** |
| Larson et al. (2022) | ASD: 34 white, 1 Latino  TD: 31 white, 2 multiracial | English | 34 ASD 34 TD | NR | 13.3 | >77 | CELF-4 CLS ≤-1.25 *SD* or RS SS ≤-1 *SD* | 111 (14) | CELF-4 SS | x |  | **9.4 (2.9) < 12.0 (1.8)** | **<.001** | **1.04** |
| Sukenik & Friedmann (2018) | NR | Hebrew | 18 ASD 90 TD | DSM-IV | 13.33 | >75 | - | NR | PETEL | x |  | **NR** | **<.0001** | **-** |
| Tyson et al. (2014) | NR | English | 44 "HFA"  34 TD | NR | 13.9 | ≥77 | - | 110.2 (12.8) | CELF-4 SS | x |  | **9.4 (3.0) < 12.0 (1.8)** | **<.01** | **0.99** |
|  |  |  |  |  |  |  |  |  |  |  | x | **9.7 (3.2) < 13.1 (1.3)** | **<.01** | **1.34** |
| **ASD = TD** | | | | | | | | | | | | | |  |
| Harper-Hill et al. (2013) | NR | English | 20 PDD 15 TD | ICD-10 | 11.58 | - | - | 99.2 (18.0) | CELF-4 SS | x |  | 9.2 (3.8) = 10.4 (2.2) | .230 | 0.4 |
|  |  |  |  |  |  |  |  |  |  |  | x | 9.5 (4.4) = 11.5 (1.8) | .289 | 0.56 |
| Manenti et al. (2023)* | NR | French | 34 ASD  41 TD | ICD-11 | 31 | - | ≤ -1.25 *SD* of the TD sample *M* on LITMUS-SR-FR | 99.1 (22.3) | LITMUS-French % | x |  | 73 (34.9) = 89.1 (9.4) | .466 | 0.66 |
| **Mixed: ASD = TD & ASD < TD** | | | | | | | | | | | | | |  |
| Georgiou & Spanoudis (2021) | "Greek native" | Greek | 16 ASD  35 TD | DSM-5 | 9.1 | ≥75 | ≤ -1.25 *SD* on ≥2 EREL subtests | 92.38 (10.6) | EREL raw score | x |  | 39.1 (17.5) = 45.9 (12.4) | .500 | 0.48 |
|  |  |  |  |  |  |  |  |  |  |  | x | **22.3 (11.7) < 29.5 (10.7)** | **<.01** | **0.65** |
| **ALI = TD** | | | | | | | | | | | | | |  |
| Riches et al. (2010) | NR | English | 16 ALI  17 TD | ICD-10 | 14.67 | ≥80 | CELF-3 core language ≤ -1.5 *SD* | 11.6 (2.4) | CELF-3 raw score | x |  | 21.2 (4.7) = 28.2 (1.5) | .538 | 1.98 |
| **ALI < ASD** | | | | | | | | | | | | | | |
| Alhassan & Marinis (2021) | NR | Arabic | 10 ALI  10 ASD | NR | 6.04 5.83 | - | Arabic PPVT ≤ -1 *SD* | 11.4 (2.2)  14.6 (1.8) | Saudi-SRT % correct | x |  | **.10 (.09) < .23 (.08)** | **.010** | **1.53** |
| **ALI = ASD** | | | | | | | | | | | | | |  |
| Georgiou & Spanoudis (2021) | "Greek native" | Greek | 24 ALI  16 ASD | DSM-5 | 6.4  9.1 | ≥75 | ≤ -1.25 *SD* on ≥2 EREL subtests | 86.9 (5.4)  92.4 (10.6) | EREL raw score | x |  | 18.3 (10.3) = 39.1 (17.5) | .070 | 1.45 |
|  |  |  |  |  |  |  |  |  |  |  | x | 9 (6.5) = 22.3 (11.7) | .980 | 1.46 |
| Whitehouse et al. (2008) | NR | English | 18 ALI  18 ASD | DSM-IV | 10.9 10.7 | ≥80 | <10th centile on ≥2: TROG, ERRNI Beach Story, TOWRE sight word & phonemic decoding, CCC-2, NEPSY NWR, NEPSY memory for sentences | 100.3 (11.7) 110.3 (14.9) | NEPSY SS | x |  | 8.4 (3.4) = 9.8 (2.7) | NR | 0.46 |
| **Group Differences Unknown** | | | | | | | | | | | | | |  |
| Botting & Conti-Ramsden (2003) | NR | English | 13 AD | DSM-IV | 10.8 | >70 | EVT < 10th centile & TROG < 50th centile | 90 (76-107) | CELF-R median % | x |  | 5 (1-9) | NR | - |
| McGregor et al. (2012) | NR | English | 12 ALI  21 ASD | NR | 10.97 | ≥85 | CELF-4 SR & FS scaled scores ≤ -1 *SD* | 101 (12.1) 113 (12.3) | CELF-4 *M* raw score | x | x | 75.5 (11.0)  137.5 (8.3) | NR | - |
| Silleresi et al. (2020) | NR | French | 16 ALI  27 ASD | DSM-5 | 8.92 | ≥80 | LITMUS SR & NWR < 77% | 92.3 (15.3) | LITMUS-French % | x |  | 31.1 (19.9)  87.9 (11.2) | NR | - |
| Manenti et al. (2023)* | NR | French | 24 ASD-Normal  10 ASD-Low | ICD-11 | 31 | - | ≤ -1.25 *SD* on LITMUS-SR-FR | 99.1 (22.3) | LITMUS-French % | x |  | **-** | **<.001** | - |
| *Note.* Significant differences in bolded text. Dx = diagnosis. LI = language impairment. SR = Sentence repetition. SP = sentence production. ASD = autism spectrum disorder. NVIQ = nonverbal intelligence quotient. *g* = Hedge’s g. TD = typically developing. ICD-10 = International Classification. "HFA" = "high functioning autism," as reported in original study. DSM-IV = Diagnostic and statistical manual of mental disorders (DSM)-4th ed. (American Psychiatric Association [APA], 1994). DSM-5 = DSM-5th ed. (APA, 2013). KORABLIK = Russian Child Language Assessment Battery (Lopukhina et al., 2019). CELF-P:2 = Clinical Evaluation of Language Fundamentals (CELF)-Preschool, 2nd Ed (Semel et al., 2004). Raw = raw score. CELF-R = CELF-Revised (Semel et al., 1987). SS = scaled score. CELF=4 = CELF-4th Ed. (Semel et al., 2003). CLS = core language score. PETEL = PETEL: A Sentence Repetition Test (Friedmann, 2000). EREL = Expressive and Receptive Language Evaluation (Spanoudis & Pathiti, 2014). ALI = autism plus language impairment. CELF=3 = CELF-3rd Ed (Semel et al., 1995). AD = autistic disorder. TROG = Test for Reception of Grammar (Bishop, 2005). ERRNI = Expression, Reception, and Recall of Narrative Instrument (Bishop, 2004). TOWRE = Test of Word Reading Efficiency (Torgesen et al., 1999). CCC-2 = Children's Communication Checklist-2nd Ed. (Bishop, 2003). NEPSY = NEPSY: A developmental neuropsychological assessment (Korkman et al., 1998). NWR = nonword repetition test. EVT = Expressive Vocabulary Test (Williams, 1997). LITMUS = The LITMUS-Sentence Repetition-French (Prévost et al., 2012). Arutiunian et al. (2022) measured NVIQ using Raven's Colored Progressive Matrices (Raven, 2000, 2004) in ASD and Kaufman Assessment Battery for Children, K-ABC II (Kaufman & Kaufman, 2004) and Wechsler Intelligence Scale for Children-Third Edition (WISC-III, 1991). In Larson et al. (2022), Landa & Goldberg (2005), and Tyson et al. (2014), NVIQ cutoff was also for FSIQ and VIQ. Riches et al. (2010) reported Wechsler Intelligence Scale for Children - III (WISC-III; Wechsler, 1992) Block Design and Picture Arrangement mean. NR = not reported. - = not possible to calculate.  *Manenti et al. (2023) reported 5 ASD participants reached the stop criterion (i.e., did not answer or answered in a fragmented way, such as by producing only one word or one phrase, for a whole block) and were excluded from analyses. Also, accuracy lower in ASD-Low than ASD-Normal (*U*(34) = 0, *p* < .001, *r* = -0.784), but means and *SD* not reported. | | | | | | | | | | | | | | |
